# Hypertension Associated Characteristics of Gut Archaea and Their Interaction Network with Bacteria

**DOI:** 10.1101/2024.04.25.24305417

**Authors:** Wen Yuan, Yihang Chen, Jian Meng, Luyun Fan, Hongjie Chi, Jing Li, Xun Zhang, Lin Feng, Pixiong Su, Jiuchang Zhong, Tianhui Li, Baicun Li, Xiaoyan Liu

**Affiliations:** Medical Research Center, Beijing Institute of Respiratory Medicine and Beijing Chao-Yang Hospital, Capital Medical University, Beijing 100020, China; Heart Center, Beijing Chao-Yang Hospital, Capital Medical University, Beijing 100020, China; Chinese Academy of Medical Sciences, Peking Union Medical College, National Key Laboratory of Cardiovascular Diseases, Fuwai Hospital, State Key Laboratory of Cardiovascular Disease, Beijing 100037, China; Department of Hypertension, Beijing Anzhen Hospital, Capital Medical University, Beijing 100029, China; Evidence-Based Medicine Center, Beijing Institute of Respiratory Medicine and Beijing Chao-Yang Hospital, Capital Medical University, Beijing 100020, China; Institute of Digestive Disease and Department of Medicine and Therapeutics, State Key Laboratory of Digestive Disease, Li Ka Shing Institute of Health Sciences, The Chinese University of Hong Kong, Hong Kong; Center of Respiratory Medicine, China-Japan Friendship Hospital, National Center for Respiratory Medicine, Institute of Respiratory Medicine, Chinese Academy of Medical Sciences, National Clinical Research Center for Respiratory Diseases, State Key Laboratory of Respiratory Health and Multimorbidity, Beijing 100029, China; Department of Respiratory and Critical Care Medicine, Beijing Institute of Respiratory Medicine and Beijing Chao-Yang Hospital, Capital Medical University, Beijing, 100020, China

**Keywords:** hypertension, gut archaeome, gut bacteria, diagnostic biomarker, gut microbiome

## Abstract

**Background:** Hypertension is a major global health challenge and a leading contributor to cardiovascular morbidity and mortality. Increasing evidence indicates that the gut microbiota contributes to the development and progression of hypertension. However, the role of gut archaea, an understudied component of the microbiome, remains poorly defined.

**Methods:** A total of 246 fecal metagenomic samples from two independent cohorts were analyzed, including 83 healthy controls and 163 individuals with hypertension. Gut archaeal composition, diversity, and interaction networks were assessed, and a diagnostic model was constructed using random forest.

**Results:** Compared with healthy controls, individuals with hypertension exhibited significantly reduced archaeal richness (Chao1 index, P = 0.0024) and distinct community structure (Adonis, P = 0.011). Hypertension status was a major host factor associated with variation in archaeal community composition. Methanobacteriota was the dominant archaeal phylum in both groups. However, key methanogenic archaea, including *Methanobrevibacter_A_smithii, Methanosphaera_sp900322125*, and *Methanomassiliicoccus_A_sp905203995*, were significantly depleted in HTN. Hypertension also altered the correlations between gut archaea and clinical indicators, including blood pressure and lipid parameters, and markedly reshaped archaeal–archaeal and archaeal–bacterial interaction networks. A diagnostic model based on three core methanogenic archaeal taxa achieved an AUC of 0.945 in the combined cohort and demonstrated strong performance in the two independent cohorts (AUC = 0.858 and 0.999).

**Conclusions:** This study provides the first comprehensive characterization of gut archaeal dysbiosis in hypertension. Reduced methanogenic archaea and remodeled cross-domain networks are hallmarks of hypertension. Gut archaeal signatures serve as promising noninvasive biomarkers.

---

Hypertension remains a major global health challenge and is strongly associated with increased risks of cardiovascular disease, stroke, and mortality.^1^ The development and progression of hypertension are influenced by a complex interplay of genetic predisposition, lifestyle behaviors, and environmental exposures.^2^ In recent years, the gut microbiome has emerged as an important contributor to systemic blood pressure regulation, attracting increasing attention for its role in hypertension pathophysiology.^3–6^ Microbial-derived metabolites are key mediators linking gut microbiota dysbiosis to cardiovascular disease severity, with trimethylamine N-oxide and imidazole propionate representing well-characterized examples.^17^

The gut microbiome comprises bacteria, archaea, viruses, and fungi and plays a central role in host metabolism, immune regulation, and overall health.^7,8^ Archaea represent a distinct and diverse domain of life alongside bacteria and eukaryotes.^9^ Although bacterial components of the microbiome have been extensively investigated, the archaeal fraction offers valuable insights into the complexity of hypertension. Recent multi-kingdom microbiome studies indicate that non-bacterial microbial taxa, including archaea and fungi, contribute to disease pathogenesis. Distinct archaeome and mycobiome signatures have been identified in conditions such as colorectal cancer and inflammatory bowel disease.^18^

Emerging studies further highlight links between gut archaea and human health.^10^ Kim et al. reported a high abundance of halophilic archaea in the gut archaeome of Korean individuals.^11^ Barnett et al. demonstrated that *Methanobrevibacter smithii* (MSS) is inversely associated with childhood asthma risk, with higher relative abundance correlating with lower disease incidence.^12^ In colorectal cancer, halophilic archaea are enriched while methanogenic archaea are depleted, with the halophilic archaeon *Natrinema sp.* J7-2 progressively increasing from healthy individuals to adenoma and colorectal cancer.^13^ Alterations in the gut archaeal community have also been reported in idiopathic pulmonary arterial hypertension (IPAH), where multi-kingdom microbial signatures including archaeal taxa provide improved diagnostic performance compared with single-kingdom markers.^34^ Despite these observations, the relationship between gut archaea and hypertension remains largely undefined.

The present study aimed to characterize gut archaeal profiles in hypertensive and normotensive individuals and to identify archaeal community alterations associated with hypertension. Using high-resolution metagenomic sequencing and integrative analytical approaches in two independent cohorts comprising 246 participants from different geographic regions,^14^ we examined archaeal community composition, diversity, and interactions with gut bacteria. In addition, we evaluated the potential diagnostic value of archaeal signatures for hypertension, providing new insights into the role of the gut microbiome in blood pressure regulation.

## Methods

### Study Cohorts

Two independent cohorts were included in this study. Cohort 1 was derived from our previously published investigation conducted in Tangshan and included both hypertensive patients and healthy individuals.^3^ The metagenomic sequencing data for this cohort are available from the European Nucleotide Archive (ENA) under BioProject accession number PRJEB13870. ^3^ Cohort 2 comprised participants from Dalian, China, including hypertensive patients and healthy controls; fecal metagenomic sequencing data for this cohort were obtained from the ENA under accession number PRJEB21612.^5^ As all participants were drawn from these previously published studies, which had obtained ethical approval from their respective institutional review boards, the present study involved no additional human subject recruitment or data collection and was therefore exempt from further ethical review.

In both cohorts, healthy controls were defined as individuals not receiving antihypertensive treatment with systolic blood pressure (SBP) ≤120 mmHg and diastolic blood pressure (DBP) ≤80 mmHg. Hypertension (HTN) was defined according to the 2025 American College of Cardiology/American Heart Association (ACC/AHA) guidelines as ≥130 mmHg or DBP ≥80 mmHg in the absence of antihypertensive medication.^35^

### Data Preprocessing and Microbial Taxonomic Analysis

Data preprocessing and microbial taxonomic profiling were performed as previously described.^33^ Metagenomic datasets obtained from the ENA public database and in-house sequencing were quality filtered using KneadData v0.10.0 with default parameters to remove host-derived sequences and low-quality reads. To improve archaeal taxonomic resolution, a custom Kraken–GTDB reference database was constructed using Genome Taxonomy Database (GTDB) version 214 genomes.^14^ To minimize sequence misclassification, fungal and viral genomes from the NCBI RefSeq database were also incorporated.^15^ The final database included 394,932 genomes representing 80,789 bacterial species, 7,777 genomes representing 4,416 archaeal species, 9,346 viral reference genomes, and 1,094 fungal reference genomes. Shotgun metagenomic reads were classified using Kraken2 v2.1.2beta with a confidence threshold of 0.05.^16^ Species abundances at each taxonomic level were subsequently re-estimated using Bracken v2.5.3 with default parameters (k=35, l=60). Archaeal and bacterial abundance profiles were then extracted separately for downstream analyses.

### Diversity and Structural Analysis of Gut Archaeal Communities

Community diversity was assessed using α-diversity and β-diversity metrics based on archaeal species abundance. Community richness and evenness were evaluated using the Chao1 and Shannon indices, respectively, and intergroup differences were tested using the Wilcoxon rank-sum test. β-diversity was examined using principal coordinates analysis (PCoA) based on Bray–Curtis distance to visualize differences in archaeal community structure between HC and HTN individuals. Statistical significance of community differences was evaluated using Adonis (Permutational Multivariate Analysis of Variance, PERMANOVA) with 999 permutations. PERMANOVA was also applied to assess the marginal explanatory power of clinical variables, including study cohort, hypertension status, age, sex, body mass index (BMI), and glycolipid metabolic indicators, on archaeal community variation.

### Archaeal and Bacterial Enterotype Analysis

Unsupervised clustering using Partitioning Around Medoids (PAM) was applied to species-level relative abundance profiles to classify archaeal and bacterial enterotypes. The optimal number of clusters was determined using the Calinski–Harabasz (CH) index. Participants were ultimately categorized into two archaeal enterotypes (Ar-C1 and Ar-C2) and two bacterial enterotypes (ET-B and ET-P).

Wilcoxon rank-sum tests were used to identify dominant characteristic species within each enterotype. Chi-square tests were performed to evaluate differences in archaeal enterotype distribution between the HC and HTN groups and to examine associations between archaeal and bacterial enterotypes.

### Differential Species Analysis

To identify gut archaeal and bacterial taxa associated with hypertension, relative abundance data were subjected to arcsine square root (AST) transformation and total sum scaling (TSS) normalization. Multivariate Association with Linear Models (MaAsLin2) was applied to detect taxa with consistent trends across both cohorts, prevalence >10%, and significant associations with hypertension after adjustment for age, sex, BMI, and cohort effects. Wilcoxon rank-sum tests were performed to confirm intergroup abundance differences of key differential species.

### Correlation and Interaction Network Analysis

Correlation matrix analyses between gut archaeal taxa and clinical variables, including age, sex, BMI, SBP, DBP, total cholesterol (TC), triglycerides (TG), low-density lipoprotein (LDL), and fasting blood glucose (FBG), were performed using OmicStudio (https://www.omicstudio.cn/tool). Differential Gene Correlation Analysis (DGCA) was used to construct archaeal–archaeal and archaeal–bacterial interaction networks separately for HC and HTN groups and to identify associations with significantly strengthened or weakened interactions in HTN. Networks were visualized using Cytoscape.

### Co-Association Analysis Between Gut Archaeal and Bacterial Communities

Associations between gut archaeal and bacterial communities were evaluated through two complementary analyses. Pearson correlation analysis was used to assess relationships between α-diversity indices (richness and Shannon index) of archaeal and bacterial communities within HC and HTN groups. Structural concordance between archaeal and bacterial communities was assessed by pairing their PCoA results using Procrustes analysis based on Bray-Curtis distances, with statistical significance evaluated using the Mantel test.

### Construction and Validation of a Hypertension Diagnostic Model Based on Gut Archaeal Features

The Boruta algorithm was applied to identify “Confirmed features” among species-level differential archaeal taxa in the combined cohort. A random forest model was subsequently used to select key features based on the mean decrease in Gini index (MeanDecreaseGini > 5). A classification model was constructed using the screened biomarkers. Model performance was evaluated using 10-fold cross-validation, with receiver operating characteristic (ROC) curves plotted and the area under the curve (AUC) calculated. Model generalizability was evaluated independently in Cohort 1 and Cohort 2.

### Statistical Analysis

All statistical analyses were performed using R software (version 4.1.0). Normally distributed continuous variables were reported as mean ± standard deviation and compared using the t-test. Non-normally distributed variables were expressed as median (interquartile range) and compared using the Wilcoxon rank-sum test. Categorical variables were summarized as number (percentage) and analyzed using the chi-square test or Fisher’s exact test. Correlations were assessed using Spearman’s rank correlation. All statistical tests were two-sided, and *P* < 0.05 was considered statistically significant.

## Results

### Clinical Baseline Characteristics of the Study Cohorts

Two independent cohorts were analyzed: Cohort 1 from a previous study conducted in Tangshan,^3^ and Cohort 2 from Dalian, China, with fecal metagenomic data obtained from the European Nucleotide Archive (ENA; accession PRJEB21612).^5^ Across all samples, the relative abundance of archaea ranged from 0.06% to 0.25%. To ensure data quality and analytical reliability, stringent filtering criteria were applied,^33^ excluding samples with missing data, <1 million aligned reads, insufficient archaeal sequencing depth (<800 archaeal reads), or abnormally elevated archaeal abundance (>1%, indicative of potential contamination) (Figure 1B).

**Figure 1.**
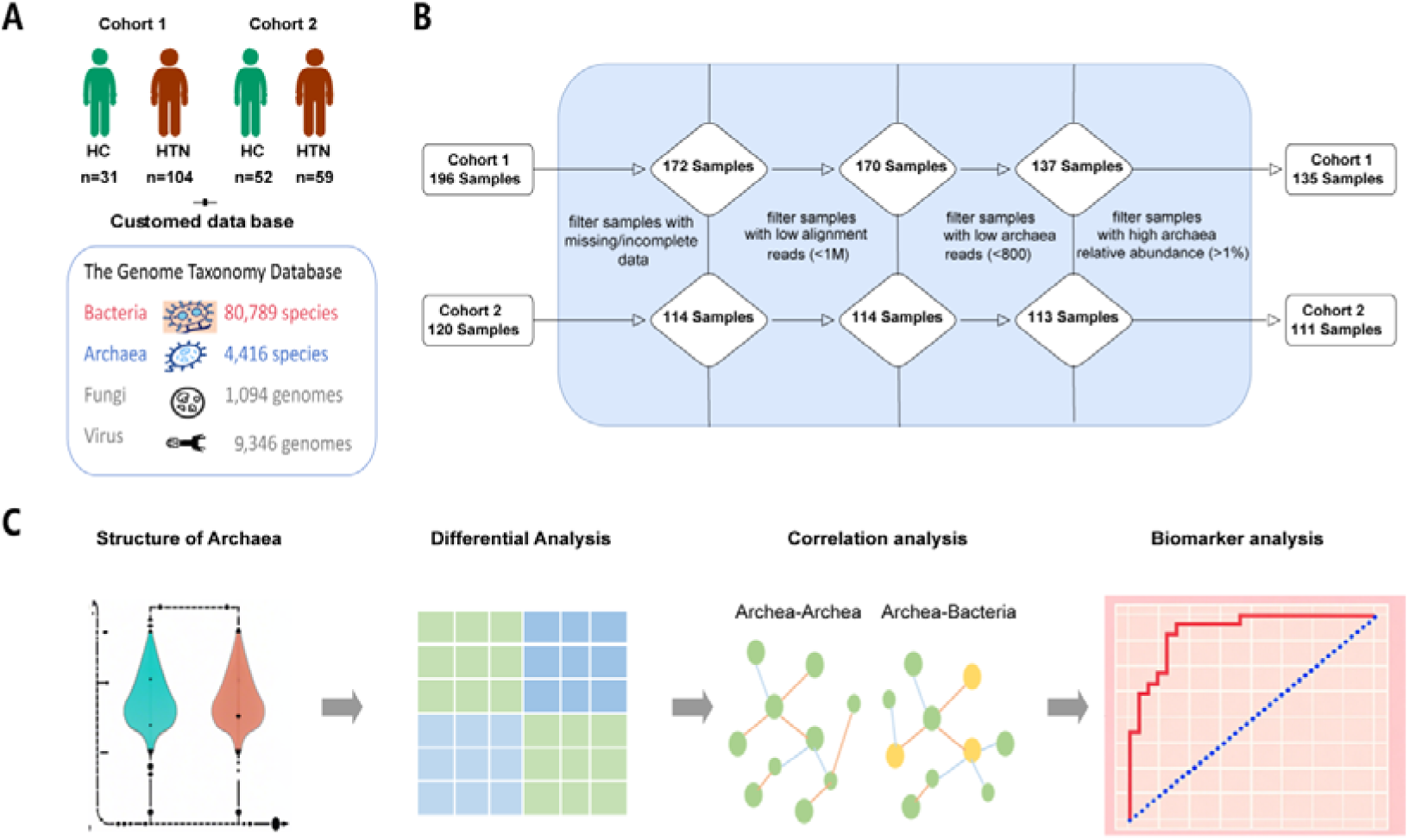
Study design and analytical workflow of gut archaeome analysis in hypertension. **A**. Study cohorts and reference databases. Cohort 1 included 31 healthy controls (HC) and 104 hypertensive patients (HTN), while Cohort 2 comprised 52 HC and 59 HTN. The integrated reference database included 80,789 bacterial species, 4,416 archaeal species, 1,094 fungal genomes, and 9,346 viral genomes. **B.** Sample screening process. Following stringent quality control procedures, 135 samples were retained from the initial 196 samples in Cohort 1, and 111 samples were retained from the initial 120 samples in Cohort 2. **C.** Analytical workflow. Bioinformatic and statistical analyses included archaeal community profiling, differential abundance analysis, interspecies co-occurrence network analysis, and biomarker identification. HC, healthy control; HTN, hypertension.

Following quality control, 135 samples were retained in Cohort 1 (HC: n = 31; HTN: n = 104) and 111 samples in Cohort 2 (HC: n = 52; HTN: n = 59) (Figures 1A–B). The combined cohort comprised 83 HC and 163 individuals with HTN (Table 1).

**Table 1.** Clinical characteristics of the study population. Normally distributed variables are presented as mean ± standard deviation, and non-normally distributed variables are expressed as median (interquartile range). HC, healthy control; HTN, hypertension; BMI, body mass index; SBP, systolic blood pressure; DBP, diastolic blood pressure; FBG, fasting blood glucose; LDL, low-density lipoprotein; TG, triglyceride; TC, total cholesterol.

| Cohort | Cohort 1 |  |  | Cohort 2 |  |  | All |  |  |
| --- | --- | --- | --- | --- | --- | --- | --- | --- | --- |
|  | HC | HTN | P value | HC | HTN | P value | HC | HTN | P value |
| sample size | 31 | 104 | N/A | 52 | 59 | N/A | 83 | 163 | N/A |
| Male ( % ) | 24 ( 77.4% ) | 98 ( 94.2% ) | < 0.01 | 29 ( 55.8% ) | 34 ( 57.6% ) | 0.844 | 53 ( 63.9% ) | 132 ( 81.0% ) | < 0.01 |
| Age | 54.45±6.07 | 53.06±5.9 | 0.255 | 56.06±8.519 | 57.17±9.642 | 0.524 | 55.46±7.696 | 54.55±7.713 | 0.381 |
| BMI | 25.620±3.581 | 25.76±3.313 | 0.839 | 23.43±2.708 | 23.61±2.893 | 0.732 | 24.25±3.224 | 24.98±3.324 | 0.098 |
| SBP | 115.4±7.948 | 142.0±17.35 | <0.001 | 111.1±6.427 | 163.6±20.95 | <0.001 | 112.7±7.290 | 149.8±21.37 | <0.001 |
| DBP | 74.59±5.674 | 90.62±10.14 | <0.001 | 71.19±6.854 | 100.2±11.77 | <0.001 | 72.46±6.613 | 94.09±11.08 | <0.001 |
| FBG | 5.380±0.939 | 5.666±1.302 | 0.275 | 6.208±2.105 | 6.476±2.432 | 0.539 | 5.912±1.815 | 5.976±1.854 | 0.798 |
| LDL | 2.680± | 2.856± | 0.620 | 2.996± | 3.064± | 0.622 | 2.888± | 2.932± | 0.806 |
|  | HC | HTN | P value | HC | HTN | P value | HC | HTN | P value |
|  | 0.787 | 1.792 |  | 0.696 | 0.756 |  | 0.739 | 1.5 |  |
| TG | 1.454±<br>0.876 | 1.819±<br>1.847 | 0.305 | 1.485±<br>0.699 | 1.852±<br>0.851 | 0.016 | 1.474±<br>0.762 | 1.831±<br>1.556 | 0.054 |
| TC | 5.490±<br>0.930 | 5.537±<br>2.388 | 0.911 | 4.973±<br>0.988 | 5.051±<br>0.984 | 0.678 | 5.158±<br>0.993 | 5.360±<br>2.004 | 0.394 |

In Cohort 1, age and BMI did not differ significantly between groups (*P* > 0.05), whereas the proportion of men was higher in HTN than in HC (94.2% vs. 77.4%, *P* < 0.01). In Cohort 2, no significant differences were observed in age, sex distribution, or BMI (*P* > 0.05). As expected, SBP and DBP were significantly elevated in HTN compared with HC in both cohorts (*P* < 0.0001). Among metabolic parameters, triglyceride (TG) levels were higher in HTN than HC only in Cohort 2 (*P* = 0.0155), whereas fasting blood glucose (FBG), low-density lipoprotein (LDL), and total cholesterol (TC) did not differ significantly between groups in either cohort (*P* > 0.05). In the combined cohort, SBP, DBP, and male proportion remained significantly higher in HTN (*P* < 0.01), with no intergroup differences in age, BMI, FBG, LDL, or TC (*P* > 0.05) (Table 1).

### Composition of the Gut Archaeal Community in Healthy and Hypertensive Individuals

Gut archaeal community composition was characterized at the phylum, order, and species levels in the combined cohort (HC = 83; HTN = 163) to define dominant taxa and intergroup differences (Figure S1).

At the phylum level, Methanobacteriota, Thermoplasmatota, and Halobacteriota predominated in both groups (Figure S1A), with Methanobacteriota as the most abundant phylum (HC: 48.73%; HTN: 40.00%).

At the order level, Methanobacteriales was dominant (HC: 48.18%; HTN: 39.61%), followed by Methanomassiliicoccales (HC: 10.52%; HTN: 11.00%). Other prevalent orders included Halobacteriales, Nitrososphaerales, and Woesearchaeales, without consistent intergroup trends (Figure S1B).

At the species level, *Methanobrevibacter_A_sp900766745* was the most abundant taxon in both groups. In HC, the next most abundant species were *Methanobrevibacter_A_smithii* and *UBA71_sp006954465*, whereas in HTN they were *UBA71_sp006954465* and *Nitrosotenuis_sp021824845*. Notably, *Methanobrevibacter_A_smithii* was markedly reduced in HTN (4.17%), whereas *Nitrosotenuis_sp021824845* increased from 1.53% in HC to higher levels in HTN. The predominance of methanogenic and halophilic archaea in both groups suggests important functional roles for these taxa within the gut microbial ecosystem and their close association with host physiological status.

### Diversity and Structural Characteristics of the Gut Archaeal Community in Hypertensive Patients

To characterize differences in gut archaeal diversity between hypertensive patients and healthy individuals, α-diversity and β-diversity analyses were performed in the combined cohort, and permutational multivariate analysis of variance (PERMANOVA) was applied to evaluate the contribution of clinical variables to community variation (Figures 2; Figures S2).

**Figure 2.**
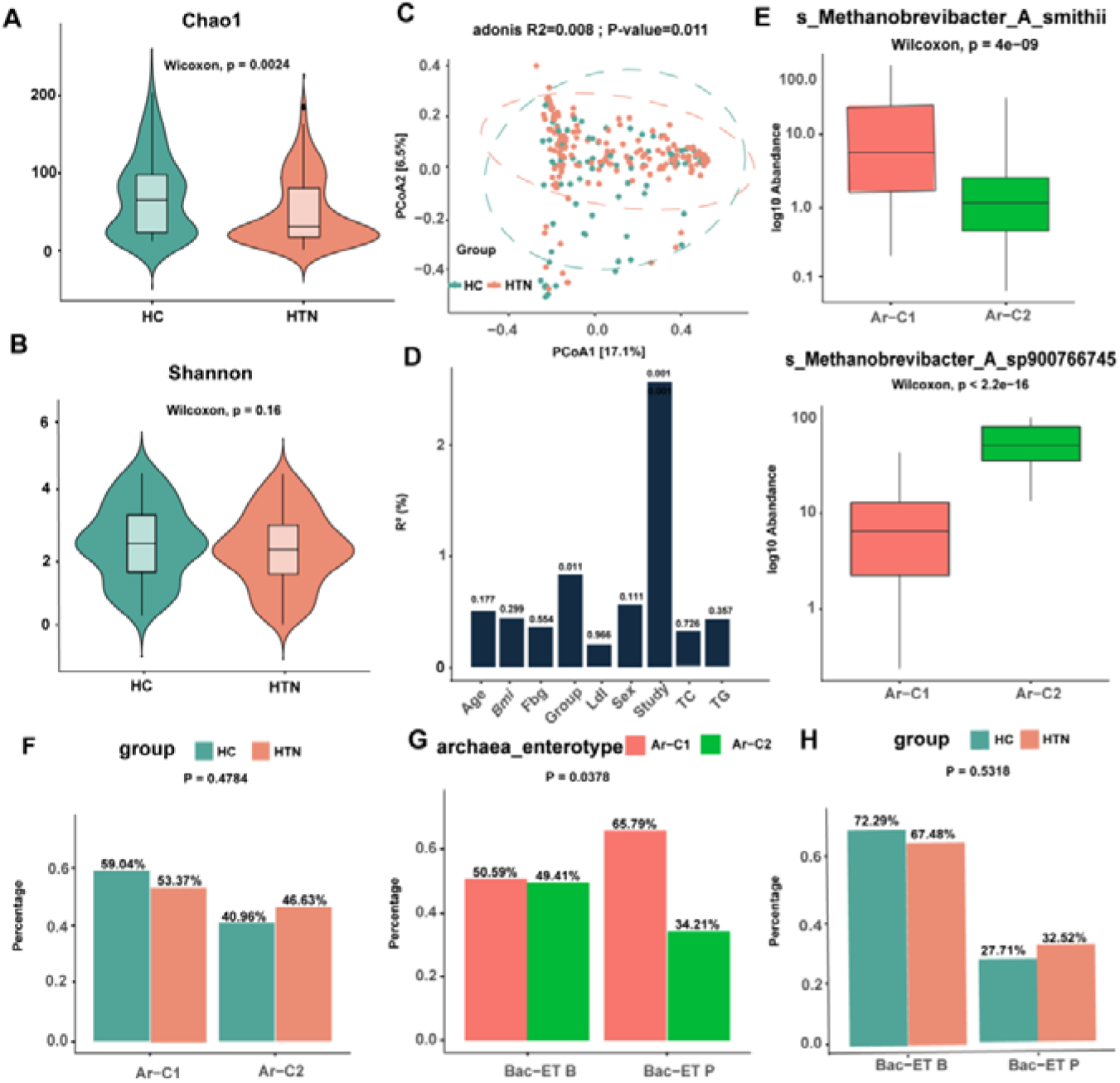
Diversity and structural characteristics of the gut archaeal community. A. Chao1 index of the gut archaeal community (Wilcoxon rank-sum test). **B.** Shannon index of the gut archaeal community (Wilcoxon rank-sum test). **C.** Principal coordinates analysis (PCoA) based on Bray–Curtis dissimilarity (Adonis test, *R*² = 0.008, *P* = 0.011). **D.** Marginal explanatory power of clinical variables on archaeal community variation assessed by PERMANOVA. **E.** Intergroup differences in the relative abundance of two representative archaeal species (Wilcoxon rank-sum test). **F.** Distribution of archaeal enterotypes (Ar-C1, Ar-C2) in the HC and HTN groups (*P* = 0.0378). **G.** Distribution of bacterial enterotypes (Bac-ET B, Bac-ET P) across archaeal enterotypes (Ar-C1, Ar-C2) (*P* = 0.5318 for Ar-C1; *P* = 0.4784 for Ar-C2). **H.** Distribution of bacterial enterotypes (Bac-ET B, Bac-ET P) in the HC and HTN groups. HC, healthy control; HTN, hypertension; PCoA, principal coordinates analysis.

α-diversity analysis demonstrated a significant reduction in archaeal richness in the HTN group compared with HC, as reflected by a lower Chao1 index (Wilcoxon test, *P* = 0.0024; Figure 2A), whereas community evenness (Shannon index) did not differ significantly between groups (*P* > 0.05; Figure 2B). These findings indicate that hypertension is associated with reduced species richness without affecting evenness. β-diversity analysis using PCoA based on Bray–Curtis distance revealed a clear separation trend in archaeal community structure between HC and HTN groups (Adonis test, *P* = 0.011; Figure 2C).

PERMANOVA analysis further showed that study cohort exerted the strongest effect on archaeal community variation (*P* = 0.001), followed by hypertension status (*P* = 0.011), whereas age, BMI, sex, and glycolipid metabolic indicators did not demonstrate significant explanatory power (*P* > 0.05) (Figure 2D). Collectively, these results indicate that hypertension is a key host factor associated with reduced archaeal richness and altered community structure.

### Distribution of Archaeal Enterotypes and Their Association with Bacterial Enterotypes

To investigate archaeal enterotype distribution and its relationship with bacterial community structure, PAM clustering was performed based on species-level archaeal abundance profiles, classifying subjects into two archaeal enterotypes: Ar-C1 and Ar-C2 (Figure S2A). Ar-C1 was dominated by *Methanobrevibacter_A_smithii*, whereas Ar-C2 was characterized by *Methanobrevibacter_A_sp900766745* (Figure 2E). Comparative analysis showed that Ar-C1 was slightly more prevalent in HC, whereas Ar-C2 was relatively enriched in HTN; however, these differences did not reach statistical significance (Figure 2F).

Using a similar approach, participants were stratified into two bacterial enterotypes: ET-B and ET-P (Figure S2B). ET-B was dominated by *Bacteroides_uniformis* and *Phocaeicola_vulgatus* (Figures S2C–D), whereas ET-P was characterized by *Prevotella_copri_B*, *Prevotella_sp021636625*, and *Prevotella_sp900551275* (Figures S2E–G). Notably, the archaeal Ar-C1 enterotype was more prevalent within the bacterial ET-P enterotype (Figure 2G). No significant differences in the distribution of ET-B and ET-P were observed between HTN and HC groups (Figure 2H). These findings suggest a close association between archaeal and bacterial enterotype structures.

### Identification of Hypertension-Associated Differential Gut Archaeal Species

To identify archaeal taxa associated with hypertension, relative abundance data from both individual and combined cohorts were subjected to arcsine square root transformation (AST) and TSS normalization. Multivariate Association with Linear Models (MaAsLin2) was applied after adjusting for age, sex, BMI, and cohort effects. A total of 13 archaeal taxa meeting the criteria of consistent trends across cohorts, prevalence >10%, and significant association with hypertension (*P* < 0.05) were identified (Figure 3A), including 4 species-level and 9 genus-level taxa. Among these, several taxa were significantly depleted in the HTN group, including *Methanosphaera_sp900322125*, *Methanobrevibacter_A_smithii*, *Methanomassiliicoccus_A_sp905203995*, *JABXJT01_sp016840125*, *JABXJT01*, *Methanococcoides*, *Methanoregula*, and *Methanosphaera*. By contrast, taxa enriched in HTN included *Methanobacterium_D*, *PALSA-986*, *UBA184*, *UBA460*, and *18H4-34*.

**Figure 3.**
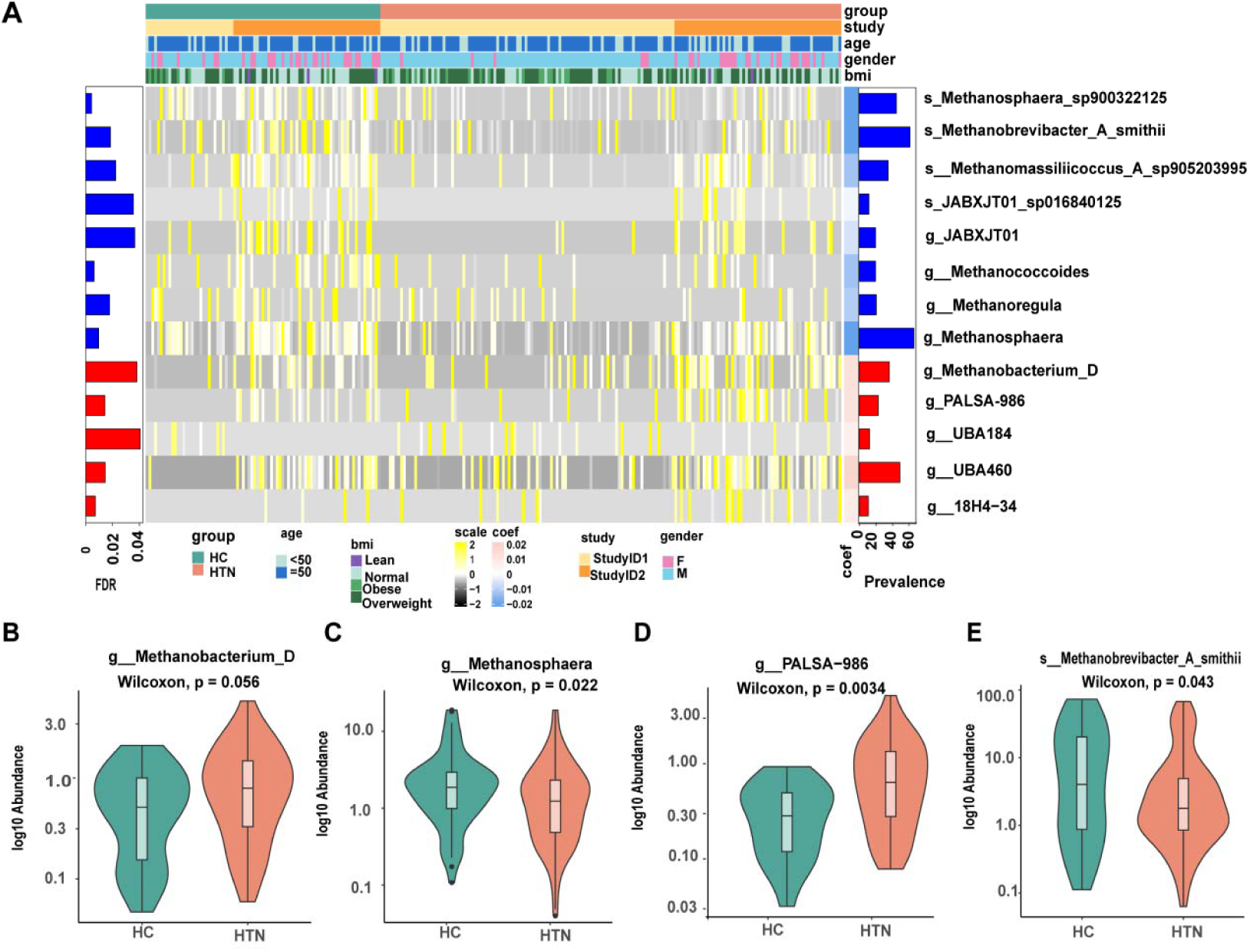
Differential gut archaeal taxa associated with hypertension. A. Heatmap of archaeal taxa identified by MaAsLin2 after adjustment for confounders (age, sex, BMI, cohort). **B–E**. Log10-transformed relative abundance comparisons of key taxa between HC and HTN groups (Wilcoxon rank-sum test): **B.** *g Methanobacterium_D* (*P* = 0.056); **C.** *g Methanosphaera* (*P* = 0.043); **D.** *g PALSA-986* (*P* = 0.022); **E.** *s Methanobrevibacter_A_smithii* (*P* = 0.0034). HC, healthy control; HTN, hypertension.

Wilcoxon rank-sum tests further supported these findings, showing increased trends for *Methanobacterium_D* (*P* = 0.056) and *PALSA-986* (*P* = 0.022), and significantly reduced abundances of *Methanosphaera* (*P* = 0.043) and *Methanobrevibacter_A_smithii* (*P* = 0.0034) in HTN compared with HC (Figures 3B–E). These results confirm that gut archaeal composition is altered in hypertension, with core differential taxa predominantly belonging to methanogenic archaea.

### Correlation Between Gut Archaea and Clinical Indicators, and Archaeal–Archaeal Interaction Network Analysis

To investigate associations between gut archaea and host clinical parameters, 41 archaeal species with prevalence >10% and consistent abundance trends across both cohorts were selected for analysis (Table S1). Spearman correlation analysis was performed between these taxa and clinical variables, including age, sex, BMI, SBP, DBP, TC, TG, LDL, and FBG (Figure 4A). Distinct correlation patterns were observed between the HC and HTN groups: In the HC group, most archaeal taxa exhibited weak correlations with blood pressure indices (|ρ| < 0.2), with only a subset showing moderate associations with age and BMI (0.2 < |ρ| < 0.4). By contrast, in the HTN group, correlations between archaeal taxa and blood pressure parameters were markedly strengthened, with several taxa demonstrating significant positive or negative associations with SBP and DBP (*P* < 0.05). Similarly, correlations between archaeal taxa and lipid parameters (TG, TC, LDL) were more pronounced in the HTN group than in the HC group. These findings indicate that hypertension is associated with a substantial reconfiguration of the relationships between gut archaea and host metabolic and hemodynamic variables.

**Figure 4.**
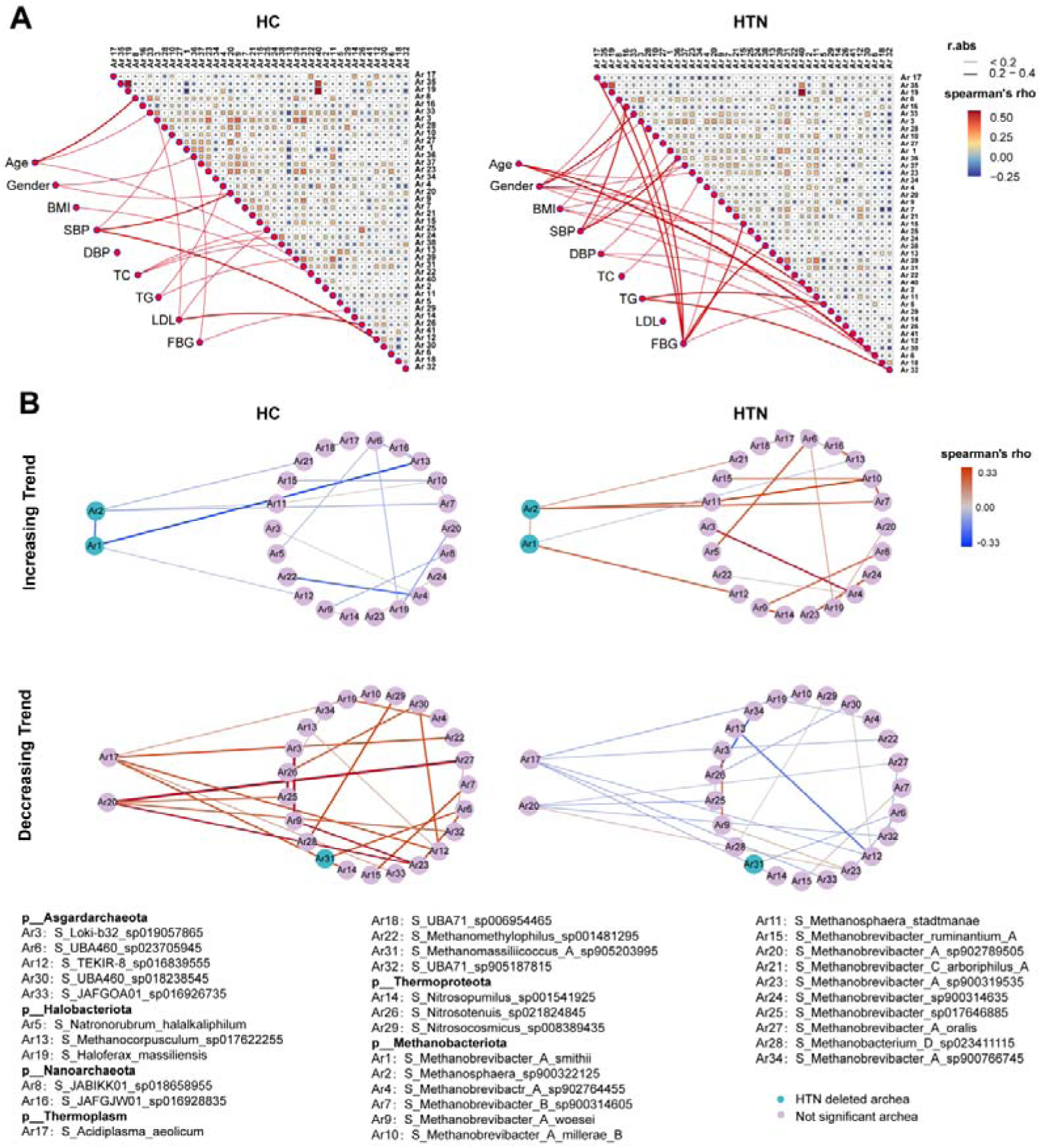
Correlation between gut archaea and clinical indicators and archaeal co-occurrence networks. **A.** Heatmap of Spearman correlations between archaeal taxa and clinical indicators, and pairwise archaeal correlations (left: HC; right: HTN). Red indicates positive correlations, blue indicates negative correlations, and line thickness reflects correlation strength. **B.** Archaeal co-occurrence network analysis stratified by abundance trends in HTN relative to HC (upper: increased taxa; lower: decreased taxa; left: HC; right: HTN). Nodes represent archaeal species (Ar1–Ar34), and edges indicate significant correlations. HC, healthy control; HTN, hypertension.

Archaeal–archaeal interaction networks constructed using DGCA further demonstrated marked alterations in network structure in the HTN group. Based on changes in association strength, the network was partitioned into modules of enhanced and weakened interactions (Figure 4B).

Within the enhanced interaction module, *Methanobrevibacter_A_smithii* and *Methanosphaera_sp900322125* occupied central positions. In the HTN group, correlations between *Methanobrevibacter_A_smithii* and *Methanosphaera_sp900322125*, *Methanocorpusculum_sp017622255*, and *TEKIR-8_sp016839555* were significantly strengthened. Similarly, *Methanosphaera_sp900322125* exhibited enhanced associations with *Methanobrevibacter_A_smithii*, *Methanobrevibacter_sp900314635*, *Methanosphaera_stadtmanae*, and *Methanobrevibacter_B_sp900314605.* By contrast, the weakened interaction module was centered on *Acidiplasma_aeolicum* and *Methanobrevibacter_A_sp002789505*. In the HTN group, correlations between *Acidiplasma_aeolicum* and *Methanobrevibacter_A_millerae_B*, *TEKIR-8_sp016839555*, and *Nitrosopumilus_sp001541925* were significantly reduced. Likewise, associations between *Methanobrevibacter_A_sp902789505* and *Methanobrevibacter_A_oralis*, *Methanobrevibacter_sp017646885*, *UBA71_sp905187815*, and *Methanobrevibacter_A_sp900319535* were significantly weakened (Figure 4B).

### Association Between Gut Archaeal and Bacterial Community Diversity and Structure

To further characterize interactions between gut archaeal and bacterial communities under hypertension, correlations between diversity indices (richness and Shannon index) were assessed, and structural concordance was evaluated using Procrustes analysis combined with the Mantel test (Figure S3).

A strong positive correlation between archaeal richness and bacterial richness was observed in both the HC group (R = 0.83, *P* < 2.2e−16) and the HTN group (R = 0.77, *P* < 2.2e−16) (Figures S3A–B). By contrast, no significant correlation was detected between archaeal and bacterial Shannon indices in either the HC group (R = 0.067, *P* = 0.55) or the HTN group (R = −0.11, *P* = 0.18) (Figures S3C–D). These findings indicate a consistent positive association between archaeal and bacterial richness irrespective of disease status. Procrustes analysis, supported by Mantel testing, demonstrated significant concordance between archaeal and bacterial community structures in both the HC and HTN groups (Figures S3E–F), further supporting coordinated ecological relationships between these microbial domains within the gut ecosystem.

### Correlation Between Gut Archaea and Bacteria, and Archaeal–Bacterial Interaction Network Analysis Under Hypertension Status

To evaluate the impact of hypertension on archaeal–bacterial interspecific associations, correlation analyses were performed using 41 archaeal species and 41 bacterial species with consistent abundance trends across both cohorts. Significant differences were observed in archaeal–bacterial correlation patterns (|ρ| > 0.2, *P* < 0.05) between the HTN and HC groups. The HC group was characterized predominantly by negative correlations, whereas the HTN group exhibited a predominance of positive correlations (Figure 5A), indicating a marked shift in interdomain association patterns under hypertensive conditions.

**Figure 5.**
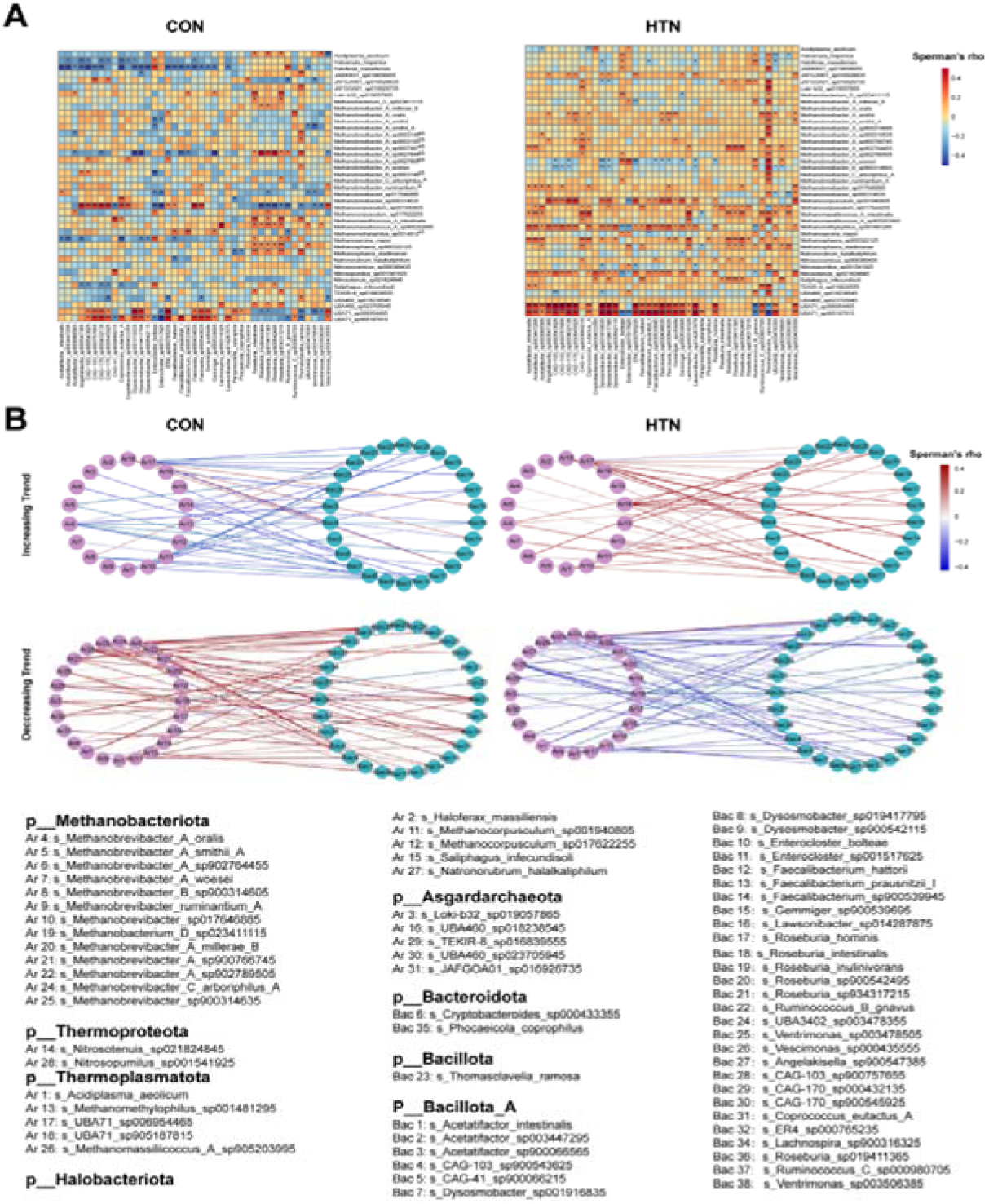
Archaeal–bacterial correlations and co-occurrence networks in hypertension. **A.** Spearman correlation heatmap of core archaeal and bacterial taxa (left: HC; right: HTN). Red indicates positive correlations, blue indicates negative correlations, and color intensity reflects correlation strength. **B.** Archaeon–bacterial co-occurrence networks stratified by abundance trends in HTN relative to HC (upper: increased taxa; lower: decreased taxa; left: HC; right: HTN). Cyan nodes represent archaeal taxa, purple nodes represent bacterial taxa, and edge colors denote the direction and magnitude of correlations. HC, healthy control; HTN, hypertension.

DGCA was subsequently applied to identify archaeal–bacterial associations with significantly altered interaction strength in the HTN group. Within the enhanced interaction network, *Nitrosotenuis_sp021824845* and *UBA71_sp006954465* emerged as central nodes, each demonstrating significantly strengthened interactions with multiple bacterial taxa compared with the HC group. Specifically, *Nitrosotenuis_sp021824845* showed enhanced associations with *Dysosmobacter_sp019417795*, *Dysosmobacter_sp900542115*, *Faecalibacterium_sp900539945*, *Gemmiger_sp900539695*, *Lawsonibacter_sp014287875*, *Roseburia_hominis*, *UBA3402_sp003478355*, and *Vescimonas_sp000435555*. Similarly, *UBA71_sp006954465* exhibited strengthened correlations with *CAG-103_sp900543625*, *Faecalibacterium_hattorii*, *Faecalibacterium_sp900539945*, *Lawsonibacter_sp014287875*, *Roseburia_intestinalis*, *Roseburia_sp934317215*, *UBA3402_sp003478355*, and *Ventrimonas_sp003478505* (Figure 5B).

By contrast, within the weakened interaction network, *Methanobrevibacter_B_sp900314605* was identified as a central archaeal node. In the HTN group, its interactions with *Angelakisella_sp900547385*, *CAG-103_sp900543625*, *CAG-103_sp900757655*, *CAG-170_sp000432135*, *CAG-170_sp900545925*, *Dysosmobacter_sp001916835*, *ER4_sp000765235*, *Faecousia_sp000434635*, and *Lawsonibacter_sp014287875* were significantly attenuated relative to the HC group (Figure 5B).

### Construction and Validation of a Hypertension Diagnostic Model Based on Gut Archaeal Features

To identify archaeal biomarkers capable of distinguishing hypertensive individuals from healthy controls, the Boruta algorithm was applied to species-level differential archaeal taxa in the combined cohort to select “Confirmed features” (Figure 6A). A random forest model was subsequently used to refine feature selection based on the mean decrease in Gini index (MeanDecreaseGini > 5). This approach identified three archaeal species as the optimal feature set: *Methanobrevibacter_A_smithii*, *Methanosphaera_sp900322125*, and *Methanomassiliicoccus_A_sp905203995*.

**Figure 6.**
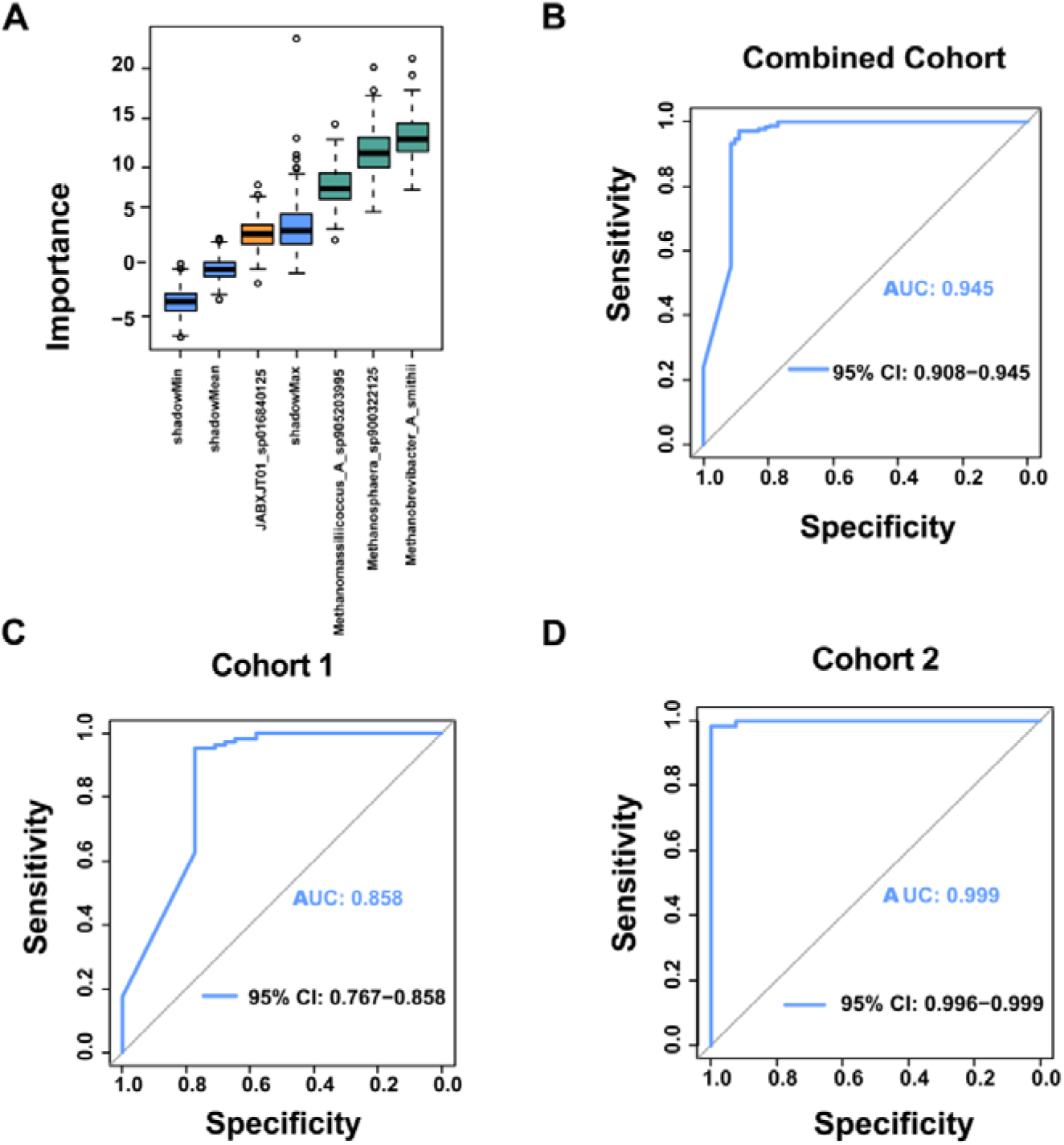
Construction and validation of a hypertension diagnostic model based on archaeal taxa. **A.** Feature importance ranking of key archaeal taxa in the random forest model. **B.** Receiver operating characteristic (ROC) curve of the model in the combined cohort (AUC = 0.945; 95% CI, 0.908–0.945). **C.** External validation in Cohort 1 (AUC = 0.858; 95% CI, 0.767–0.858). **D.** External validation in Cohort 2 (AUC = 0.999; 95% CI, 0.996–0.999). AUC, area under the curve; CI, confidence interval; ROC, receiver operating characteristic.

A classification model constructed using these three taxa demonstrated strong discriminative performance, achieving an area under the receiver operating characteristic curve (AUC) of 0.945 (95% CI, 0.908–0.945) in the combined cohort (Figure 6B). External validation confirmed robust generalizability, with an AUC of 0.858 (95% CI, 0.767–0.858) in Cohort 1 and 0.999 (95% CI, 0.996–0.999) in Cohort 2 (Figures 6C and D). These findings indicate that the identified archaeal signature provides a stable and effective means of distinguishing hypertension status across independent populations and supports its potential utility as a diagnostic biomarker.

## Discussion

This study provides a comprehensive characterization of the gut archaeal community in hypertension, integrating compositional, diversity, and network-level analyses across two independent cohorts comprising 246 participants. In addition, a diagnostic model based on archaeal species was established. The findings demonstrate that hypertension is associated with marked alterations in the gut archaeal community, particularly reductions in methanogenic archaea, which exhibit close associations with host blood pressure parameters. These results suggest a potential contributory role of gut archaea in the pathogenesis and progression of hypertension.

Archaea represent essential yet underexplored constituents of the human gut microbiota.^19–21^ Although many archaeal taxa are adapted to extreme environments,^22^ a subset colonizes moderate ecological niches within the human body, including the gastrointestinal tract, skin, respiratory tract, and genitourinary system.^23,24^ Progress in this field has historically been constrained by the limited culturability of archaeal species. However, advances in high-throughput sequencing and metagenomic approaches now enable detailed interrogation of archaeal community structure and function within complex microbial ecosystems.^10,13,14,25,26^

Consistent with prior reports, Methanobacteriota was identified as the dominant archaeal phylum in the gut.^13^ Diversity analyses revealed a significant reduction in archaeal richness (Chao1 index) in the HTN group relative to the HC group, whereas community evenness (Shannon index) remained unchanged. These findings indicate that hypertension primarily affects species richness rather than distributional uniformity within the archaeal community, a pattern also observed in colorectal cancer.^33^ PERMANOVA further demonstrated that hypertension status was a major contributor to archaeal community variation, second only to cohort-related effects, underscoring its role as a key driver of archaeal community remodeling.

Differential abundance analyses identified multiple archaeal taxa associated with hypertension. After adjustment for confounding variables, several methanogenic archaea at both the genus and species levels were significantly depleted in the HTN group, including *Methanococcoides*, *Methanoregula*, *Methanosphaera*, *Methanobrevibacter_A_smithii*, *Methanosphaera_sp900322125*, and *Methanomassiliicoccus_A_sp905203995*. By contrast, several uncultured candidate taxa, including *PALSA-986*, *UBA184*, and *UBA460*, were enriched in the HTN group. Methanogenic archaea function as primary hydrogen consumers in the gut and contribute to metabolic homeostasis through syntrophic interactions with hydrogen-producing bacteria.^31,32^ Their depletion may result in hydrogen accumulation, with downstream effects on short-chain fatty acid (SCFA) production and intestinal barrier integrity, thereby influencing blood pressure regulation.^28,29^ This mechanism is consistent with the broader paradigm of microbiota-derived metabolites mediating cardiovascular risk, wherein microbial dysbiosis promotes the generation of pro-hypertensive metabolites.^17^

Enterotype analysis further revealed distinct archaeal community configurations. The Ar-C1 enterotype, dominated by *Methanobrevibacter_A_smithii,* was more prevalent in the HC group, whereas the Ar-C2 enterotype, dominated by *Methanobrevibacter_A_sp900766745,* was relatively enriched in the HTN group, although the distribution difference did not reach statistical significance. Notably, archaeal enterotypes exhibited significant associations with bacterial enterotypes, with Ar-C1 preferentially linked to the ET-P bacterial enterotype. Complementary Procrustes and Mantel analyses demonstrated a strong positive correlation in species richness and high concordance in community structure between archaeal and bacterial communities in both the HC and HTN groups. These findings highlight the tightly coupled and potentially coevolved relationships between archaeal and bacterial domains within the gut ecosystem, consistent with multi-kingdom microbiome studies demonstrating coordinated interdomain interactions in disease contexts.^18^

Further network-based analyses demonstrated that hypertension profoundly reconfigures both archaeal–archaeal and archaeal–bacterial interaction patterns. In the HC group, archaeal taxa exhibited only weak associations with clinical blood pressure parameters, and archaeal–bacterial interactions were predominantly characterized by negative correlations. By contrast, in the HTN group, correlations between archaeal taxa and clinical indicators, including blood pressure and lipid profiles, were markedly strengthened, accompanied by a shift toward predominantly positive archaeal–bacterial interactions. Notably, interaction strengths between *Nitrosotenuis_sp021824845* and *UBA71_sp006954465* and canonical SCFA-producing genera, including *Faecalibacterium* and *Roseburia,* were significantly increased in the HTN group. Conversely, interactions between *Methanobrevibacter_B_sp900314605* and multiple commensal bacterial taxa were significantly attenuated.^27,30^ These findings suggest that hypertension is associated not only with compositional alterations in the archaeal community but also with disruption of the coordinated ecological balance between archaea and bacteria. Such network-level remodeling may contribute to intestinal microecological instability and promote disease progression.

This study further establishes, for the first time, a hypertension diagnostic model based on archaeal features. Three key archaeal species identified through Boruta feature selection and random forest modeling—*Methanobrevibacter_A_smithii*, *Methanosphaera_sp900322125*, and *Methanomassiliicoccus_A_sp905203995*—demonstrated strong discriminative performance in the combined cohort (AUC = 0.945), with consistent generalizability across two independent cohorts (AUC = 0.858 and 0.999, respectively). These results highlight the potential of archaeal taxa, as non-bacterial components of the microbiome, to serve as robust biomarkers for hypertension, particularly in the context of metabolic-related chronic diseases.

Several limitations warrant consideration. First, this study is observational in design; although findings were validated across two independent cohorts, causal relationships between archaeal alterations and hypertension cannot be established. Future studies incorporating in vivo functional approaches, such as fecal microbiota transplantation in germ-free models and targeted colonization with specific methanogenic archaea, are required to elucidate causal mechanisms and assess the direct impact of archaeal taxa on host blood pressure regulation. Second, all participants were derived from Chinese populations, limiting the generalizability of these findings. Validation in multi-ethnic, large-scale prospective cohorts is therefore necessary. Third, the present analysis focused on taxonomic composition based on metagenomic sequencing and did not address functional gene content, metabolic pathways, or active metabolite profiles. Integration of multi-omics approaches, including metatranscriptomics, metabolomics, and metaproteomics, will be essential to further delineate the mechanistic pathways linking gut archaea to host physiology.

Importantly, the complexity of causal inference must be acknowledged. Although robust associations between gut archaea and hypertension were identified, the directionality of this relationship remains unresolved. It is unclear whether archaeal dysbiosis contributes to hypertension development or arises as a consequence of the hypertensive state. Longitudinal cohort studies and interventional trials targeting the gut microbiota will be critical to clarify these relationships and define potential therapeutic implications.

In summary, this study systematically delineates the dysbiotic features of the gut archaeal community in hypertension, identifies a consistent depletion of methanogenic archaea, including *Methanobrevibacter_A_smithii,* and demonstrates extensive remodeling of archaeal–bacterial interaction networks. In addition, a high-performing, non-invasive diagnostic model based on gut archaeal biomarkers was established. These findings expand current understanding of the gut microecological contributions to hypertension and provide a foundation for the development of microbiome-based strategies for early detection, risk stratification, and targeted intervention. Continued advances in anaerobic archaeal isolation and functional characterization, together with deeper investigation of host–microbe interactions, are expected to further position gut archaea as a promising frontier in cardiovascular metabolic disease research.

## Data Availability

Metagenomic sequences for cohort 1 and cohort 2 are accessible through the European Nucleotide Archive (ENA) under the identifiers PRJEB13870 and PRJEB21612, respectively. Other data produced in the present study are available upon reasonable request to the authors.

https://www.ebi.ac.uk/ena/browser/view/PRJEB13870?show=reads

https://www.ebi.ac.uk/ena/browser/view/PRJEB21612

## Acknowledgments

We thank Phoebe Chi, MD, from Liwen Bianji (Edanz) (www.liwenbianji.cn), for editing a draft of this manuscript.We thank Edanz for providing English language editing support during the preparation of this manuscript.

## What Is New?

1. This study provides the first comprehensive metagenomic characterization of the human gut archaeome in hypertension.
2. Hypertension is associated with reduced archaeal richness, depletion of key methanogenic archaea, and extensive remodeling of archaeal–bacterial networks.
3. A 3-archaeon biomarker panel shows high diagnostic accuracy in two independent cohorts.

## What Is Relevant?

1. Gut bacteria contribute to blood pressure regulation, but the role of archaea in hypertension remains largely unexplored.
2. Methanogenic archaea maintain gut metabolic homeostasis; their depletion may disrupt microbial balance and promote hypertension.
3. Cross-domain interactions between archaea and bacteria are markedly perturbed in hypertension.

## Clinical Implications

1. Gut archaeal signatures represent promising noninvasive biomarkers for hypertension screening.
2. Targeted restoration of methanogenic archaea may offer a novel strategy for hypertension management.
3. These findings expand the microbial basis of hypertension and support translational applications.

## Perspectives

The present study unveils the previously unrecognized landscape of the gut archaeome in human hypertension, highlighting altered archaeal diversity, reduced methanogenic archaea, and disrupted archaeal–bacterial cross-domain interactions as key features of hypertension. These findings expand the microbial framework of blood pressure regulation beyond bacteria and underscore the potential contribution of archaea to gut metabolic homeostasis and cardiovascular health. The identified archaeal biomarker panel holds promise as a noninvasive diagnostic tool for hypertension screening in clinical settings. Further studies are warranted to verify the causal relationship between archaeal dysbiosis and blood pressure elevation, explore underlying metabolic and immunological mechanisms, and evaluate whether targeted modulation of methanogenic archaea can serve as a complementary strategy for hypertension prevention and management. Collectively, these observations open new avenues for investigating the archaeome as a novel target in cardiovascular diseases.

