## Supplementary Figures for "Hypertension Associated Characteristics of Gut Archaea and Their Interaction Network with Bacteria"

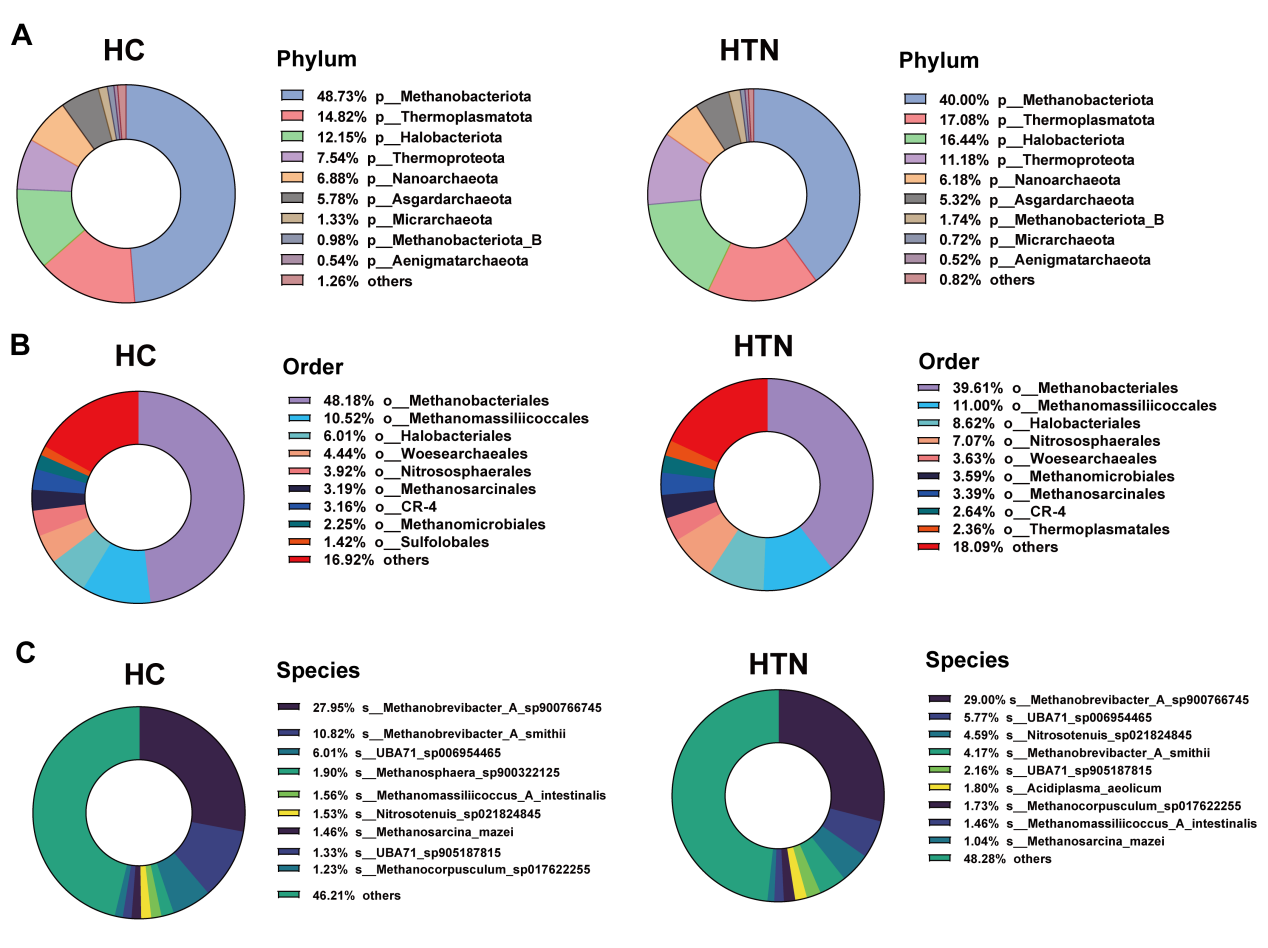

**Supplemental Figure 1. Gut archaeal community composition at the phylum, order, and species levels in the combined cohort.**

**A.** Phylum level. Methanobacteriota was the dominant phylum in both groups (HC: 48.73%; HTN: 40.00%), with an increased relative abundance of Thermoplasmatota observed in the HTN group (17.08%). **B.** Order level. Methanobacteriales was the most abundant order in both groups (HC: 48.18%; HTN: 39.61%), followed by Methanomassiliicoccales (HC: 10.52%; HTN: 11.00%). **C.** Species level. Methanobrevibacter_A_sp900766745 was the most abundant species in the HC group (27.95%), with a marked decrease in the HTN group (5.77%). By contrast, the cumulative relative abundance of other archaeal species increased in the HTN group (48.28%). HC, healthy control; HTN, hypertension.

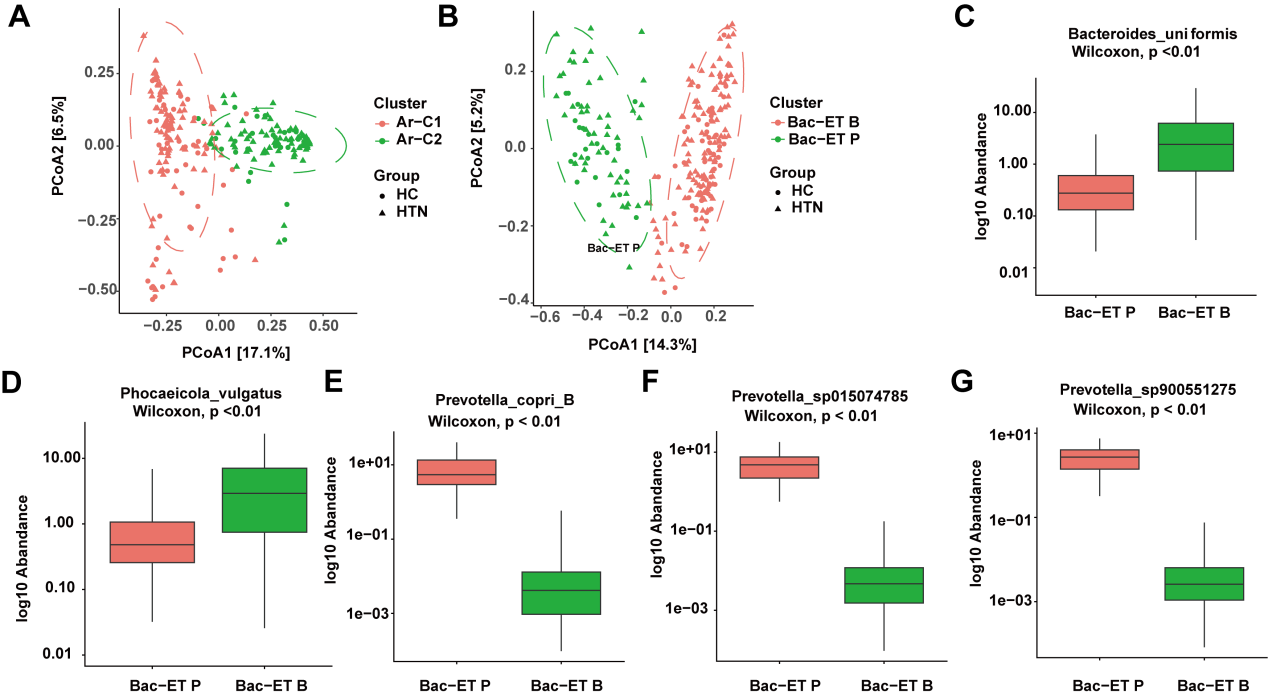

**Supplemental Figure 2. Associations between archaeal and bacterial enterotypes and differential abundance of characteristic bacterial taxa.**

**A.** PCoA based on Bray–Curtis dissimilarity illustrating archaeal enterotype distribution (Ar-C1/Ar-C2) across HC and HTN groups. **B.** PCoA illustrating bacterial enterotype distribution (Bac-ET B/Bac-ET P) across HC and HTN groups. **C–G.** Log10-transformed relative abundance comparisons of representative bacterial species between enterotypes (Wilcoxon rank-sum test): **C.** s_Phocaeicola_vulgatus; **D.** s_Bacteroides_uniformis; **E.** s_Prevotella_copri_B; **F.** s_Prevotella_sp015074785; **G.** s_Prevotella_sp900551275. HC, healthy control; HTN, hypertension; PCoA, principal coordinates analysis.

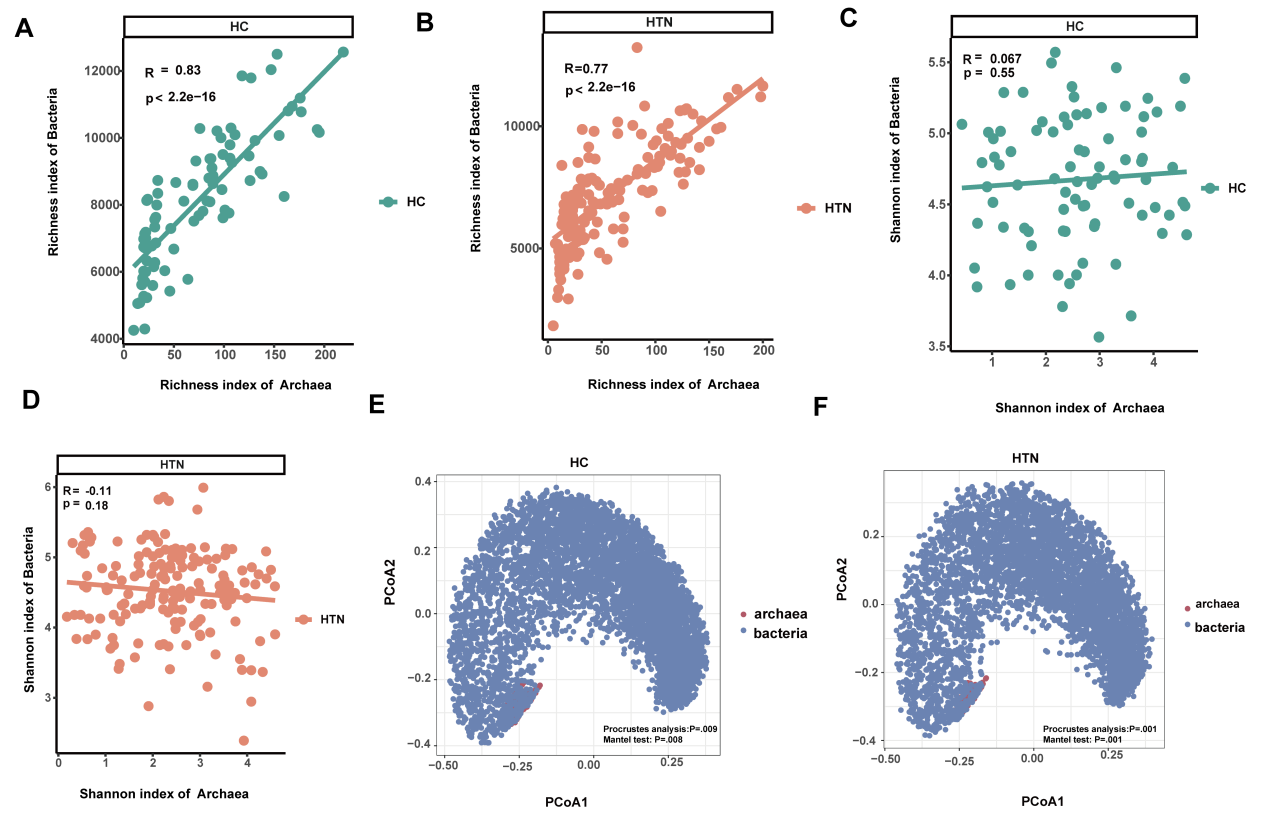

**Supplemental Figure 3. Associations between gut archaeal and bacterial diversity and community structure**

**A.** Correlation between archaeal and bacterial richness in HC (Spearman’s rho = 0.83, P < 2.2e-16). **B.** Correlation between archaeal and bacterial richness in HTN (Spearman’s rho = 0.77, P < 2.2e-16). **C.** Correlation between archaeal and bacterial Shannon index in HC (Spearman’s rho = 0.067, P = ns). **D.** Correlation between archaeal and bacterial Shannon index in HTN (Spearman’s rho = −0.11, P = 0.18). **E.** Procrustes analysis of archaeal and bacterial community structure in HC (with Mantel test). **F.** Procrustes analysis of archaeal and bacterial community structure in HTN (with Mantel test). HC, healthy control; HTN, hypertension.
